# Cribriform Status at Follow-up Biopsy Is Critical for Predicting Prostate Cancer Active Surveillance Failure in the MRI Era: a Transatlantic Cohort Study

**DOI:** 10.64898/2026.10.19.26365918

**Authors:** Nikita Sushentsev, George Xu, Irene G. Li, Son Do, Jasmine Liu, Max Kim, Christof Kastner, Sean Fernando, Anne Y. Warren, Ahmed Shabaik, Anders M. Dale, Michael Liss, Tristan Barrett, Tyler M. Seibert

**Affiliations:** Department of Radiology, Addenbrooke’s Hospital and University of Cambridge, Cambridge, United Kingdom; Department of Radiation Medicine and Applied Sciences, University of California San Diego, San Diego, California, USA; Department of Urology, Cambridge University Hospitals NHS Foundation Trust, Cambridge, United Kingdom; School of Clinical Medicine, University of Cambridge, Cambridge, United Kingdom; Department of Pathology, Cambridge University Hospitals NHS Foundation Trust, Cambridge, United Kingdom; Department of Pathology, University of California San Diego, La Jolla, CA, USA; Department of Radiology, University of California San Diego School of Medicine, La Jolla, CA, USA; Department of Urology, University of California San Diego, La Jolla, CA, USA; Department of Bioengineering, University of California San Diego Jacobs School of Engineering, La Jolla, CA, USA

**Keywords:** Prostate cancer, active surveillance, magnetic resonance imaging, cribriform carcinoma

## Abstract

**Purpose:** To assess whether follow-up MRI, biopsy status, and timing of cribriform disease detection predict active surveillance (AS) failure in a contemporary transatlantic cohort.

**Materials and Methods.:** Retrospective MRI-driven AS cohort of 619 men at two academic centers (UK/USA), 2012-2024. Primary outcome: AS failure (disease-specific mortality, metastasis, grade group [GG] ≥4 progression, or post-treatment biochemical recurrence). Key exposures: final MRI and biopsy status, timing of cribriform ≥GG2 detection. Cumulative incidence was derived; independent predictors used multivariable Fine-Gray models.

**Results.:** Over median follow-up of 6.4 years, 39/619 men (6%) experienced AS failure. Cribriform ≥GG2 was detected in 82/619 (13%) on follow-up biopsy versus 24/619 (4%) at baseline, comparably in baseline GG1 (10%) and non-cribriform GG2 (9%), and only in MRI-visible tumors. Three-year AS failure after final biopsy was 31% in men with cribriform ≥GG2 versus 0-2% with stable or non-cribriform disease; 44% of failures had stable MRI. On Fine-Gray analysis, cribriform ≥GG2 was the only independent predictor of AS failure in re-biopsied men (SHR 30; 95% CI 5.2-167; *P*<0.001); no type of MRI progression category was independently predictive. Among 595 men cribriform-negative at baseline, 6-year AS failure after a second biopsy was 34% if cribriform ≥GG2 was detected versus 7% if still cribriform-negative.

**Conclusions.:** Cribriform status at follow-up biopsy independently predicts AS failure, whereas MRI progression does not. The three-fold higher cribriform prevalence at follow-up likely reflects baseline sampling error. Because newly detected cribriform tumours were exclusively MRI-visible, early repeat biopsy of patients with MRI-visible disease may considerably improve risk stratification.

## Introduction

Active surveillance (AS) is an established management strategy for patients with low-risk and favorable intermediate-risk prostate cancer (PCa), designed to avoid or defer radical treatment while maintaining curative intent. Despite significant protocol variability, most contemporary AS pathways rely on baseline MRI and biopsy features to determine patient eligibility and guide subsequent follow-up. However, the distinct clinical value of follow-up MRI and repeat biopsies in AS patients remain controversial^1^. Challenges include subjectivity of serial MRI interpretation^2^, the negative impact of repeat biopsies on patient adherence and initial AS uptake^3^, and few multicenter studies of longitudinal AS predictors.

Our previous multicenter work established the baseline imaging and histological predictors of AS failure^4^. We showed that while MRI-driven AS is highly safe for patients with grade group 1 (GG1) and MRI-invisible, cribriform-negative GG2 disease, baseline MRI visibility and cribriform morphology are strong independent predictors of AS failure. Whereas that work defined who should enter AS, the present study addresses how patients should be monitored once enrolled. Using serial MRI and repeat-biopsy data from a transatlantic cohort, we assess whether follow-up biopsy or serial MRI predict AS failure, compare cribriform disease detection at baseline and follow-up biopsies, and derive an actionable repeat-biopsy strategy to improve risk stratification.

## Materials and methods

### Study design and cohort characteristics

This retrospective cohort study was approved by the institutional review boards of the Cambridge University Hospitals (CUH/18/3592) and University of California San Diego (191878). Men enrolled on MRI-driven AS at two academic centers between March 2012 and March 2024 were included (**Fig. 1A**); patients with less than 12 months of follow-up were excluded. The study design, patient selection criteria, baseline biopsy protocols, and AS follow-up schedules are described in our prior publication^4^ and summarized in the **Supplementary Information**. Briefly, all patients underwent baseline multiparametric MRI with targeted MRI-ultrasound fusion biopsy for MRI-visible disease and systematic biopsy otherwise. Follow-up comprised quarterly PSA monitoring, annual MRI and clinical review, and repeat biopsies either protocol-mandated at 36 months or clinically triggered by PSA kinetics or radiological progression.

**Fig. 1.**
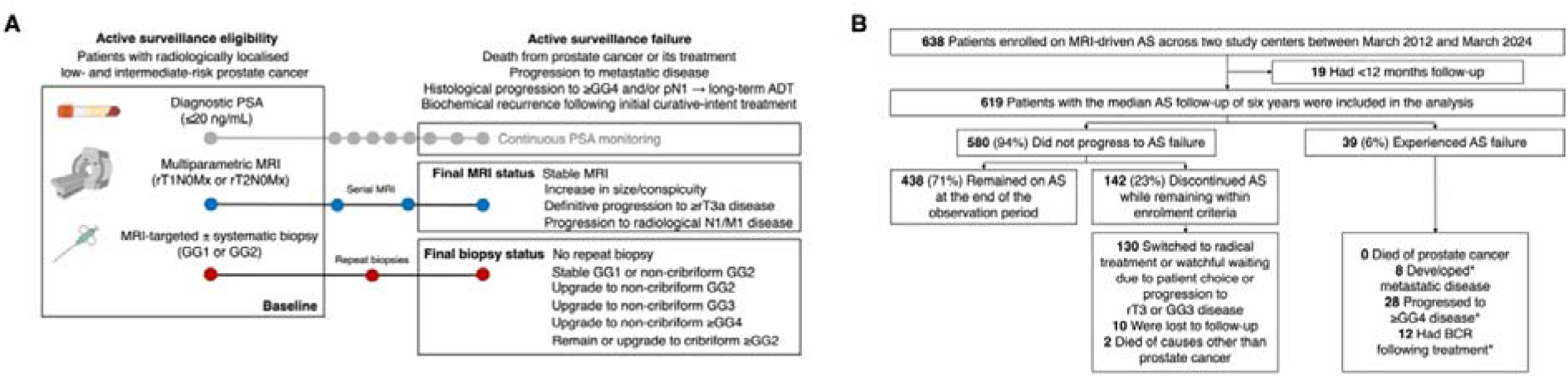
Study design and patient flow diagram. **(A)** Overview of AS eligibility criteria, baseline assessment, follow-up exposures, and AS failure definitions used in this study. **(B)** Flow diagram depicting patient enrolment, exclusions, and clinical outcomes. Asterisks denote that some patients experienced more than one AS failure outcome in differing sequence. AS, active surveillance; ADT, androgen deprivation therapy; BCR, biochemical recurrence; GG, grade group; MRI, magnetic resonance imaging; PI-RADS, Prostate Imaging–Reporting and Data System; PSA, prostate-specific antigen.

### Outcomes and exposures

The primary outcome was AS failure^4,5^, a composite of PCa-specific mortality, radiological progression to metastatic disease, histological progression to GG ≥4 disease, or biochemical recurrence following curative-intent treatment (further defined in **Supplementary Information**). Composite AS failures were further stratified by hierarchical index event as described in the **Supplementary Information**.

Key follow-up exposures were “final MRI status” and “final biopsy status”, defined as the most advanced radiological and histological finding documented across all serial assessments during the AS follow-up period (**Fig. 1A**). Final MRI status was classified as: (i) stable MRI (no significant change in size or conspicuity, no evidence of locally advanced or metastatic disease); (ii) significant increase in lesion size or conspicuity (specifically reported as such by the radiologist); (iii) definitive radiological progression to locally advanced disease (≥rT3a; gross extraprostatic extension or seminal vesicle invasion); or (iv) radiological N1/M1 disease. Final biopsy status was classified as: (i) no repeat biopsy performed; (ii) stable GG1 or non-cribriform (NC) GG2 disease; (iii) upgrade to NC GG2; (iv) upgrade to NC GG3; (v) upgrade to NC ≥GG4; or (vi) remain or upgrade to cribriform ≥GG2 disease at any point during AS.

## Statistics

The longitudinal evolution of MRI and biopsy status during AS was summarized using Sankey diagrams stratified by baseline MRI visibility (PI-RADS 1–2 *vs* 3–5) and baseline histological status (GG1 *vs* NC GG2 *vs* cribriform GG2). Cumulative incidence functions (CIFs) were estimated using the method of Kalbfleisch and Prentice, and intergroup differences were assessed using Gray’s test. Independent predictors of AS failure were assessed using multivariable Fine-Gray subdistribution hazard models adjusted for pre-specified baseline covariates (age, PSA density, GG, and PI-RADS), fitted separately for final biopsy status and final MRI status. Among patients without baseline cribriform, a landmark analysis (time origin: date of second biopsy, or two years from enrolment for those not rebiopsied) compared AS failure by second-biopsy result, and multivariable logistic regression identified baseline predictors of subsequent cribriform detection. Further details on statistical analyses are provided in **Supplementary Information**. Analyses were performed using MATLAB R2025b (MathWorks, Natick, MA) and GraphPad Prism v11.0.1. A two-sided *P* <0.05 was considered statistically significant.

## Results

### Cohort characteristics and AS outcomes

Of 638 men enrolled, 619 met the inclusion criteria (**Fig. 1B**). Of these, 580/619 (94%) did not experience AS failure over the median follow-up of 6.4 years, while 39/619 (6%) experienced AS failure, including eight (1%) cases of metastatic disease. Baseline characteristics are summarized in **Table 1**.

**Table 1.** Baseline Cohort Characteristics. AS indicates active surveillance, GG grade group, IQR interquartile range, NC non-cribriform, PI-RADS Prostate Imaging-Reporting and Data System score, PSA prostate-specific antigen. Abbreviations: AS, active surveillance, GG, grade group, IQR, interquartile range, NC, non-cribriform, PI-RADS, Prostate Imaging-Reporting and Data System, PSA, prostate-specific antigen.

|  |  |  |  |
| --- | --- | --- | --- |
| Baseline<br>characteristic | Whole<br>cohort | Patients<br>without AS | Patients with<br>AS failure |

**Table 1. Baseline Cohort Characteristics.**
|  | (n = 619) | failure<br>(n = 580) | (n = 39) |
| --- | --- | --- | --- |
| Age, yrs<br>median (IQR) | 66<br>(60-70) | 66<br>(60-70) | 66<br>(63-71) |
| AS follow-up, yrs<br>median (IQR) | 6.2<br>(3.8-8.8) | 6.4<br>(4.0-8.9) | 4.0<br>(2.8-4.6) |
| PSA, ng/mL<br>median (IQR) | 5.8<br>(4.3-7.7) | 5.7<br>(4.3-7.7) | 6.2<br>(4.3-7.0) |
| PSA density<br>median (IQR) | 0.11<br>(0.08-0.16) | 0.11<br>(0.08-0.16) | 0.11<br>(0.08-0.19) |
| PI-RADS 1-2<br>n (%) | 186<br>(30%) | 180<br>(31%) | 6<br>(15%) |
| PI-RADS 3-5<br>n (%) | 433<br>(70%) | 400<br>(69%) | 33<br>(85%) |
| GG1<br>n (%) | 467<br>(75%) | 446<br>(77%) | 21<br>(54%) |
| GG2 NC<br>n (%) | 128<br>(21%) | 116<br>(20%) | 12<br>(31%) |
| GG2 cribriform<br>n (%) | 24<br>(4%) | 18<br>(3%) | 6<br>(15%) |

### Longitudinal evolution of tumor MRI and histological characteristics

At baseline, 186/619 men (30%) had MRI-invisible (PI-RADS 1-2) and 433/619 (70%) had MRI-visible (PI-RADS 3-5) disease. Most retained their baseline visibility status at final MRI (81% of invisible, 85% of visible lesions); among men with visible disease, 39% showed radiological change (size/conspicuity increase, ≥rT3a, or N1/M1), detailed in **Fig. 2A-B** and the **Supplementary Information**.

**Fig. 2.**
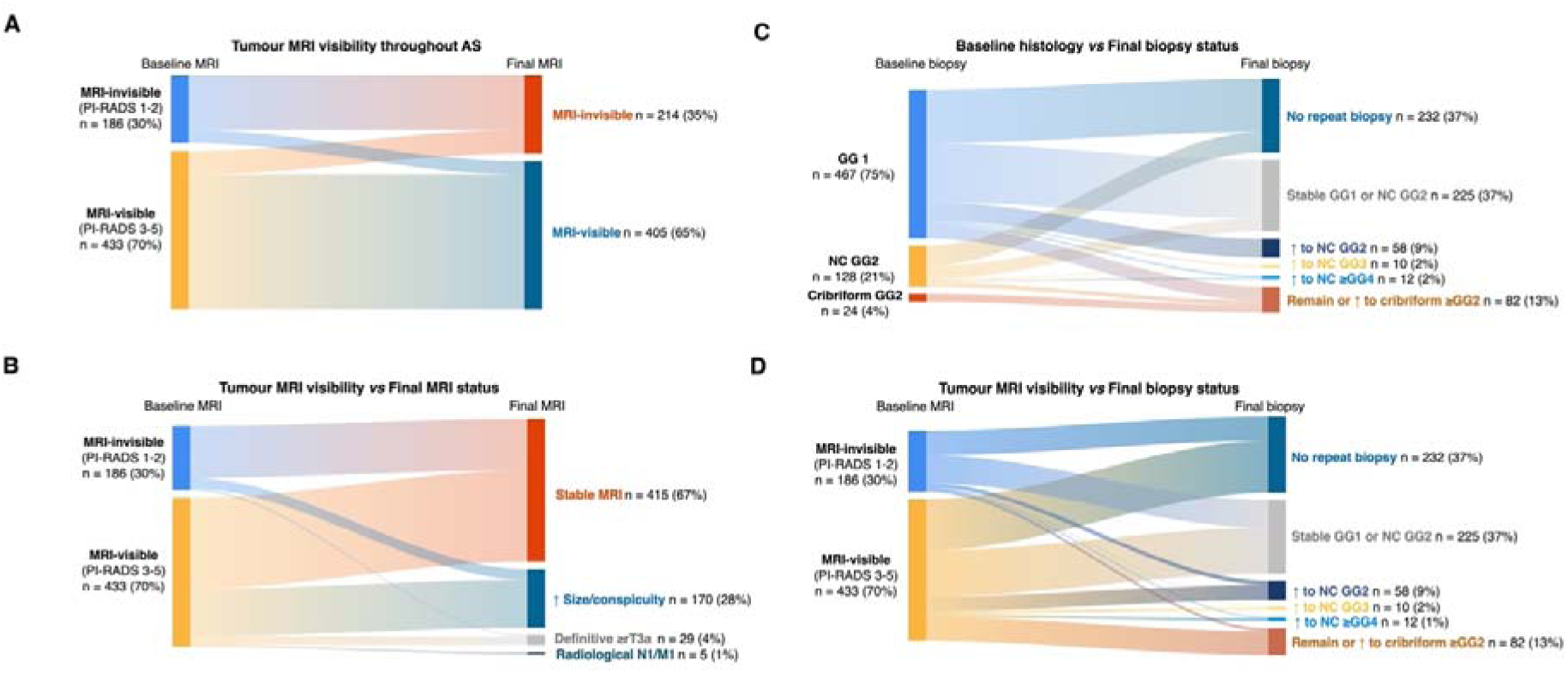
Longitudinal evolution of tumor MRI visibility and biopsy status during AS. **(A)** Sankey plot depicting the change in binary MRI visibility status between baseline and final MRI assessment. **(B)** Sankey plot illustrating the relationship between baseline MRI visibility and detailed final MRI status category. **(C)** Sankey plot depicting the relationship between baseline biopsy histological category and final biopsy status. Non-cribriform grade upgrades (to NC GG2, NC GG3, and NC ≥GG4) comprised 82 patients (13%) in total, with individual upgrade categories represented by separately coloured bands within this combined group. **(D)** Sankey plot illustrating the relationship between baseline MRI visibility and final biopsy status. Relative changes (percentages) across different groups are provided in **Supplementary Information**. AS, active surveillance; GG, grade group; MRI, magnetic resonance imaging; NC, non-cribriform; PI-RADS, Prostate Imaging–Reporting and Data System.

At baseline biopsy, 467/619 patients (75%) had GG1, 128/619 (21%) had non-cribriform (NC) GG2, and 24/619 (4%) had cribriform GG2 disease. Overall, 232/619 men (37%) did not undergo a repeat biopsy during AS; 225/619 (37%) had stable GG1 or NC GG2 disease at their final assessment; 80/619 (13%) had a cribriform-negative histological upgrade (to NC GG2, NC GG3, or NC ≥GG4); and 82/619 (13%) had cribriform ≥GG2 disease identified at any point during AS **(Fig. 2C-D)**.

This latter finding represents a three-fold increase in the detected prevalence of cribriform disease relative to the 24/619 men (4%) harboring it at baseline biopsy. Cribriform morphology was detected at comparable frequencies across histological subgroups: 10% of men with baseline GG1 and 9% of those with NC GG2 disease had cribriform identified on at least one follow-up biopsy (**Fig. 2C**). Among men with MRI-invisible disease at baseline, cribriform was subsequently detected in 5% of patients, all of whom had developed new MRI-visible lesions; cribriform disease was never detected in those who remained persistently MRI-invisible (**Fig. 2D**).

### Follow-up MRI status *vs* AS failure

Cumulative incidence curves for AS failure stratified by final MRI status are shown in **Fig. 3A**. The 3-year cumulative incidence was 5% for radiologically stable disease, 10% for size or conspicuity increase, and 7% for ≥rT3a progression; combining any MRI progression versus stable MRI yielded 3-year AS failure rates of 9% and 5%, respectively (*P*<0.01; **Supplementary Fig. S1**). Importantly, 44% of AS failures showed a stable final MRI (**Fig. 3B**).

**Fig. 3.**
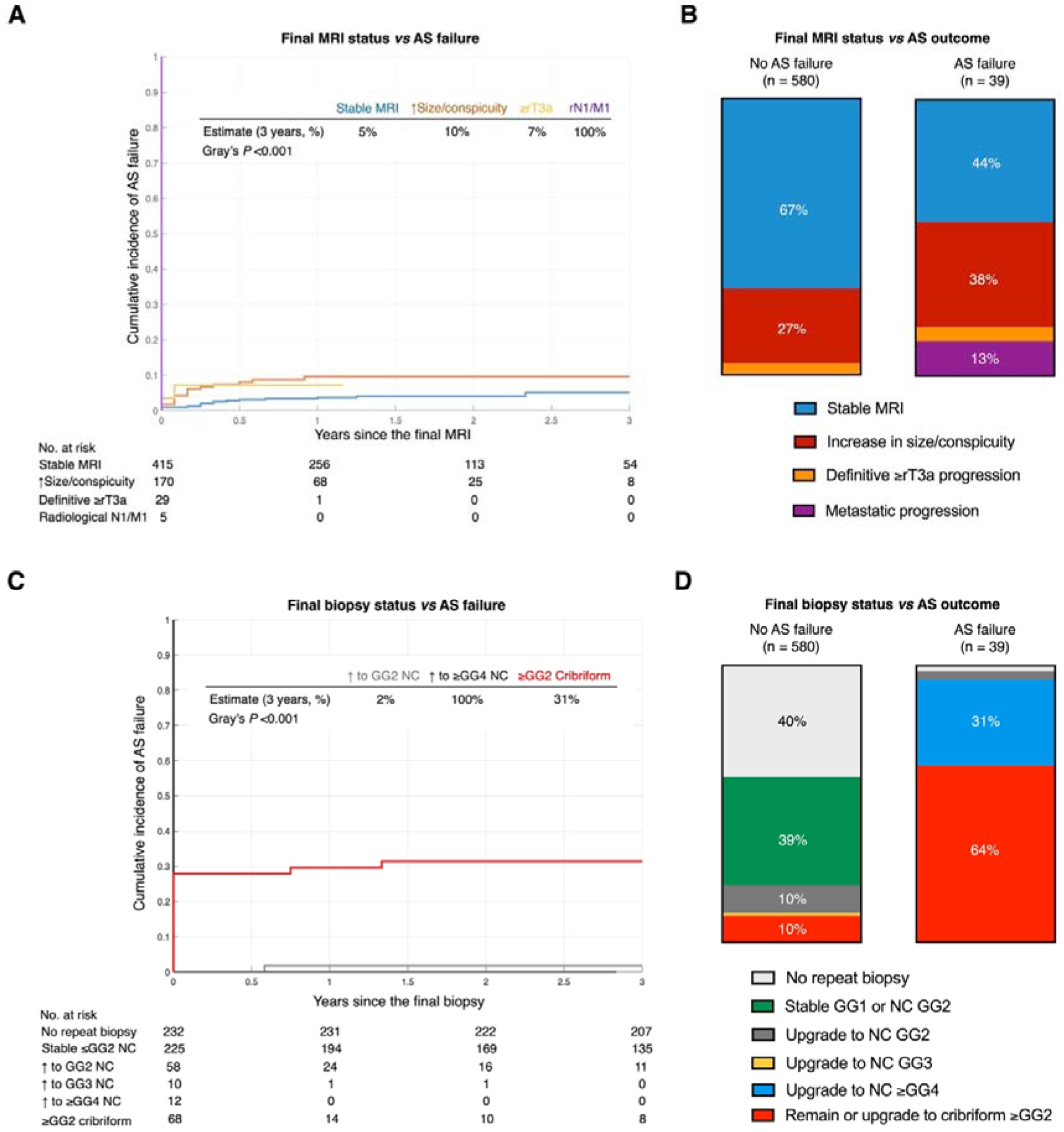
Cumulative incidence of AS failure and distribution of final MRI and biopsy status by AS outcome. **(A)** Competing-risk cumulative incidence functions for AS failure stratified by final MRI status. Three-year cumulative incidence estimates are displayed for each group. **(B)** Stacked bar charts depicting the proportional distribution of final MRI status categories among patients with and without AS failure. **(C)** Competing-risk cumulative incidence functions for AS failure stratified by final biopsy status. Three-year cumulative incidence estimates are displayed for groups with a non-zero event rate. Groups with 0% three-year cumulative incidence (no repeat biopsy; stable GG1 or NC GG2) are not individually labelled in the figure header but are shown in the at-risk table. **(D)** Stacked bar charts depicting the proportional distribution of final biopsy status categories among patients with and without AS failure. In panels (A, C), patients with radiological N1/M1 disease experienced AS failure at or near the time of staging (3-year estimate 100%), as radiological progression to metastatic disease constitutes an AS failure criterion by definition. Numbers at risk represent patients remaining event-free and without a competing risk at each time point. Beyond the three years, one patient with no repeat biopsy had AS failure (repeat biopsy not performed due to radiological progression to metastatic disease which prompted the start of androgen deprivation therapy), and two more patients with cribriform ≥GG2 disease developed AS failure; no additional AS failure events were observed in the other biopsy status groups. AS, active surveillance; BCR, biochemical recurrence; GG, grade group; MRI, magnetic resonance imaging; NC, non-cribriform.

**Fig. 4.**
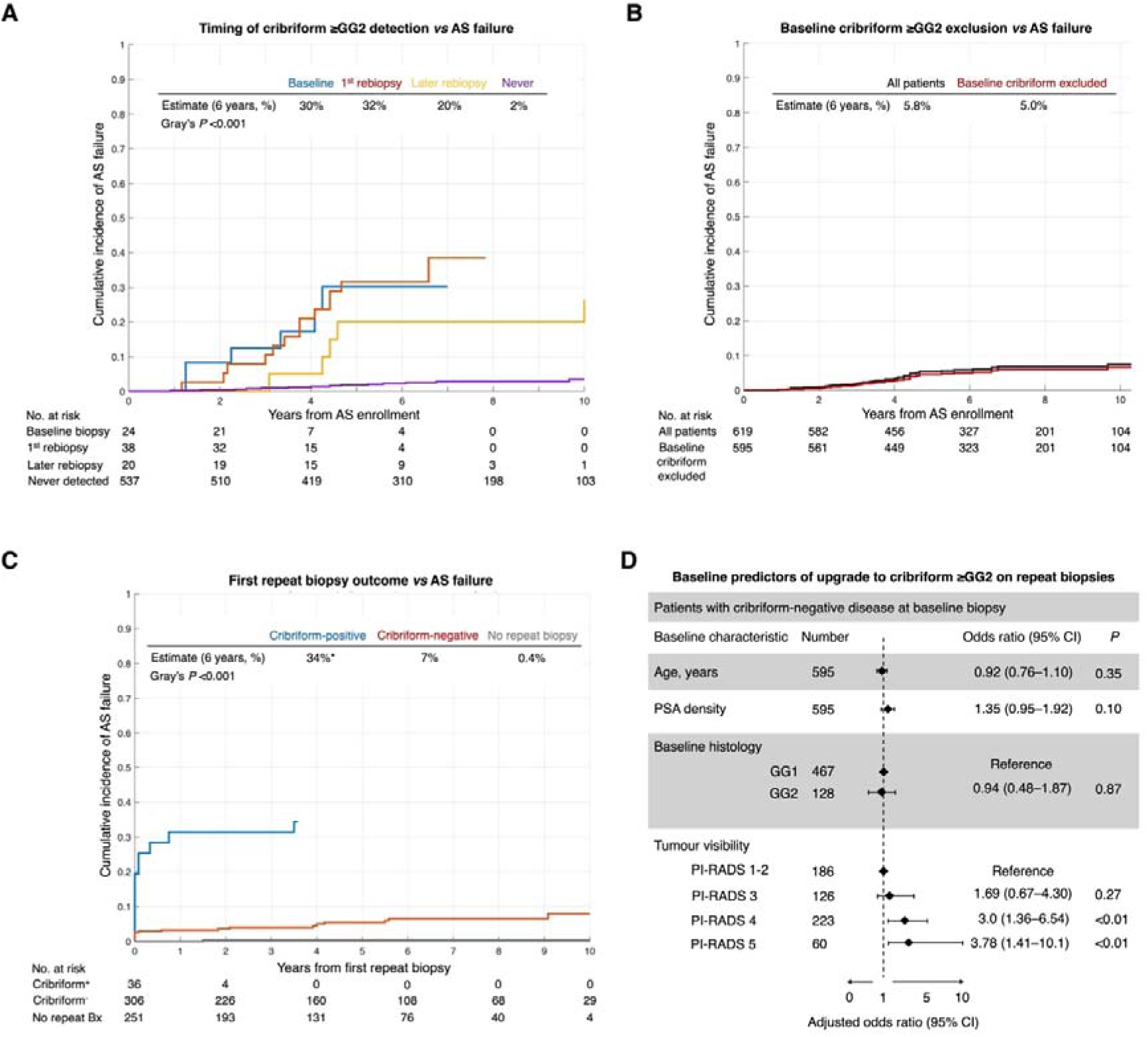
Cribriform detection during AS: timing, clinical impact, and value of the first repeat biopsy. **(A)** Competing-risk cumulative incidence functions for AS failure from the date of AS enrolment, stratified by when cribriform ≥GG2 disease was first detected: at baseline biopsy (blue), at the first repeat biopsy (orange), at a later biopsy only (yellow), or never detected during AS (purple). **(B)** Competing-risk cumulative incidence functions for AS failure from the date of AS enrolment, comparing the full cohort (black) against a hypothetical cohort excluding patients with cribriform detected at baseline biopsy only (red), modelling the real-world impact of current EAU guideline exclusion criteria. **(C)** Competing-risk landmark analysis of AS failure from the date of the first repeat biopsy among 595 patients without baseline cribriform, stratified by second biopsy result: cribriform-positive (blue), cribriform-negative (orange), or no repeat biopsy performed (grey). **(D)** Forest plot showing multivariable logistic regression results for baseline predictors of cribriform ≥GG2 detection on follow-up biopsy among 595 patients without baseline cribriform. For patients without a repeat biopsy, the landmark time origin was set to two years from AS enrolment, corresponding to the median interval to first repeat biopsy. Two patients with cribriform-positive second biopsy results experienced AS failure before the date of their second biopsy and were therefore not included in the landmark analysis at time zero; these patients are captured in the first-rebiopsy cribriform group in panel (A). *Asterisk denotes that no patients in the cribriform-positive group remained at risk beyond year 4, as all had either experienced AS failure (34%) or exited AS for treatment by this time point; the CIF estimate reflects the point of complete group depletion from surveillance. AS, active surveillance; GG, grade group; OR, odds ratio; PI-RADS, Prostate Imaging-Reporting and Data System.

### Follow-up biopsy status *vs* AS failure

Cumulative incidence curves for AS failure stratified by final biopsy status are shown in **Fig. 3C**. Men with NC ≥GG4 disease at their final biopsy experienced AS failure at or near the time of ≥GG4 detection (3-year AS failure rate 100%), as ≥GG4 progression constitutes an AS failure criterion. The 3-year AS failure rate was 0% for men with no repeat biopsy or stable NC ≤GG2 disease, and 2% for those with a NC GG2 upgrade. By contrast, the 3-year AS failure rate was 31% for men with cribriform ≥GG2 morphology at any point during AS, more than a three-fold increase relative to any radiological progression category.

Among the 39 men who experienced AS failure, 64% had cribriform morphology documented on at least one AS biopsy, compared with fewer than 10% of those who did not experience failure (**Fig. 3D**). Unlike NC GG≥4 failures, which progressed almost entirely through the GG≥4 pathway, failures in cribriform-positive men also included metastasis and post-treatment BCR (**Supplementary Fig. S2**). Of the eight patients who developed metastatic disease, five (63%) had cribriform GG≥2 disease.

On multivariable Fine-Gray analysis, detection of cribriform ≥GG2 at repeat biopsy independently predicted AS failure among rebiopsied men after adjustment for baseline age, PSA density, GG, and PI-RADS (SHR 30; 95% CI 5.2-167; P<0.001), whereas no category of MRI progression was an independent predictor in the whole cohort (**Supplementary Fig. S3**).

### Timing of cribriform detection and AS failure

Patients were stratified by when cribriform ≥GG2 disease was first identified during AS: at baseline biopsy (n=24; 4%), at the first repeat biopsy (n=38; 6%), at a later biopsy (n=20; 3%), or never detected (n=537; 87%). The 6-year cumulative incidence of AS failure was 30% for patients with baseline cribriform ≥GG2 and 32% for those with cribriform ≥GG2 first detected at the first repeat biopsy; the median interval from baseline to first repeat biopsy was 1.9 years (IQR 0.75-3.5 years). Among patients with cribriform ≥GG2 first identified only at third or later biopsy, the 6-year cumulative incidence of AS failure was 20% from baseline. Among patients in whom cribriform was never detected, the 6-year cumulative incidence of AS failure was 2% (**Fig. 5A**).

Upfront exclusion of 4% patients with baseline cribriform ≥GG2 reduced six-year cumulative incidence of AS failure from 5.8% to 5%, underscoring the limitations of a single biopsy for tumor phenotyping **(Fig. 5B)**. To estimate the added value of the first repeat biopsy, a secondary analysis was performed among 595 patients without baseline cribriform, with time zero (T0) set to the date of the second biopsy or, for those without a second biopsy, two years from AS enrollment. The 6-year cumulative incidence from T0 was 34% for patients with cribriform ≥GG2 at second biopsy (n=36), 7% for those with a negative second biopsy (n=306), and 0.4% for those without a repeat biopsy (n=251; *P*<0.001; **Fig. 5C**).

To identify which baseline characteristics independently predicted cribriform ≥GG2 detection on follow-up biopsies among patients without baseline cribriform, a multivariable logistic regression was performed. PI-RADS 4 (OR 3.0, 95% CI 1.4-6.5; *P*<0.01) and PI-RADS 5 (OR 3.8, 95% CI 1.4-10.1; *P*<0.01) were the only independent predictors of cribriform detection on follow-up biopsies; baseline GG2 (*vs* GG1), PSA density, and age (*P*>0.05 for all) were not (**Fig. 5D**; **Supplementary Fig. S3**). Median time from AS enrolment to cribriform ≥GG2 detection was 3.2 years (IQR 1.9-4.6).

## Discussion

With a 6% AS failure rate and no prostate cancer deaths over a median six-year follow-up, this study confirms that contemporary MRI-driven AS is highly safe for most men with localised PCa. These outcomes extend our prior finding that most AS failures happen in patients with cribriform disease^4^ by demonstrating that its predictive weight is not confined to baseline detection but accumulates throughout surveillance. Conversely, patients with cribriform-negative disease at repeat biopsies experience very low AS failure rates. The 15-year ProtecT trial data show that benefit from early radical treatment is largely confined to patients with cribriform morphology, whether identified at biopsy^6^ or prostatectomy^7^, reframing AS as monitoring for previously undetected cribriform morphology rather than for any MRI change or GG upgrading, since cribriform indicates treatment benefit for preventing metastasis. The clinical utility of a single repeat biopsy in achieving this goal is substantial: among patients without cribriform disease at baseline, those with cribriform disease at first repeat biopsy had a six-year AS failure rate of 34% compared with 7% for those with a negative result: a nearly five-fold difference actionable within the first two years of AS.

The three-fold increase in detected cribriform prevalence between baseline (4%) and follow-up (13%) is likely attributable to baseline sampling error rather than *de novo* disease development. Both targeted and systematic biopsies sample only a fraction of the prostate, and targeted biopsy sensitivity for cribriform status is approximately 50% against surgical pathology^8,9^, meaning that most patients identified as cribriform-positive during AS had this pattern likely undersampled at baseline. This is supported by comparable follow-up cribriform detection rates across patients classified as GG1 (10%) and cribriform-negative GG2 (9%) at baseline. However, while 95% of cribriform tumors were MRI-visible at baseline, the remaining 5% were detected only after the appearance of new MRI-visible lesions, suggesting that MRI visibility is a potential hallmark of cribriform PCa.

The finding that 44% of AS failure events occurred in men with a stable final MRI has a coherent mechanistic explanation: if the predominant predictor of failure is cribriform disease present but undetected at baseline, there is no biological imperative for that lesion to change in imaging appearance before causing systemic harm. The aggressive phenotype is pre-existing; serial MRI detects lesion growth or metastatic spread but not its histological constitution. The absolute separation between MRI-based risk categories (5% vs 9% at 3 years) contrasts sharply with the difference between stable cribriform-negative disease (0-2%) and cribriform detection (31%). Excluding patients with baseline cribriform reduces the six-year AS failure rate only marginally (5.8% to 5.0%), reflecting baseline sampling error. Proposals for MRI-only AS without scheduled or triggered repeat biopsies risk perpetuating this diagnostic blind spot for a clinically meaningful subset of patients.

This is not, however, an argument for indiscriminate repeat biopsies or “diagnostic” prostatectomies. Men with cribriform-negative disease across multiple assessments, including those upgraded to cribriform-negative GG2, have effectively zero risk of AS failure, consistent with the ∼100% specificity but ∼50% sensitivity of biopsy for cribriform^8,9^, whereby each negative targeted biopsy approximately halves the odds of undetected cribriform disease. The rational approach is risk-stratified biopsy guided by baseline MRI visibility, with repeat biopsies limited to targeted and perilesional approaches as these have been shown to be sufficient to detect most new cribriform tumors^10^. Cribriform-specific imaging^11^ and tissue-based biomarkers^5,12,13^ may augment this strategy and are undergoing clinical validation. The six-year failure rates of 7% for cribriform-negative second biopsy and 0.4% for clinically selected patients without repeat biopsy demonstrate that a single repeat assessment within two years captures most of the achievable diagnostic benefit in patients who had unequivocally MRI-visible (PI-RADS 4-5) disease at baseline. Conversely, patients with MRI-invisible or equivocal disease (PI-RADS 1-3) may avoid repeat biopsies unless developing new or progressing MRI-visible disease.

This study has several limitations. Its retrospective design and protocol variation across institutions are inherent constraints, mitigated by using broadly defined, clinically actionable exposure categories. A substantial proportion of patients (38%) did not undergo repeat biopsy, which may introduce ascertainment bias as those most likely to decline biopsy may also harbor undetected cribriform disease, thereby further underestimating its true prevalence and prognostic significance. However, the low AS failure rate in this subgroup suggests lower overall clinical risk. Generalizability to community settings may be limited by inter-reader variability in radiology and pathology reporting and by differences in biopsy technique and protocol adherence.

## Conclusions

MRI-driven AS is very safe for most men at six-year follow-up. Cribriform morphology detected at any surveillance biopsy is a strong and independent predictor of AS failure; the presence or absence of any cribriform morphology should therefore be consistently reported in all AS biopsy reports. The three-fold increase in detected cribriform prevalence during AS likely reflects sampling error at baseline, which may be significantly rectified by a single repeat biopsy performed within two years of AS enrolment in patients with PI-RADS 4-5 disease. Stability on MRI is insufficient to rule out previously undersampled cribriform cancer, and MRI progression alone is not a reliable indicator of biological aggressiveness. In practice, integrating the present results with prior literature suggests a three-step framework: patients with cribriform ≥GG2 at baseline should not commence AS; those with PI-RADS 4-5 disease at enrolment should undergo a targeted repeat biopsy within two years – if cribriform is detected, consideration of prompt curative treatment is warranted, and if not, further biopsy may be deferred unless MRI demonstrates new or unequivocally progressing lesions; and patients with PI-RADS 1-3 disease and a clinically stable course may avoid protocol-mandated rebiopsy altogether.

## Data Availability

All data produced in the present study are available upon reasonable request to the authors.

## Funding Acknowledgments

The study was funded by the National Institute for Health and Care Research (NIHR), UK, and Cancer Research UK Cambridge Centre. The views expressed are those of the authors and not necessarily those of the NIHR or the Department of Health and Social Care. N.S. acknowledges support from NIHR (Academic Clinical Fellowship) and Emmanuel College, Cambridge (Research Fellowship). A.Y.W. is supported by the Urological Malignancies Programme of the Cancer Research UK Cambridge Centre (C9685/A25177) and NIHR Cambridge Biomedical Research Centre (BRC-1215-20014). TMS acknowledges support from the Prostate Cancer Foundation (24CHAL03).

## Supplementary Information

### Supplementary Methods

#### AS description

Upon active surveillance (AS) enrollment, all patients underwent a baseline multiparametric MRI (mpMRI). Individuals lacking MRI-visible lesions received a standard 12-core systematic transrectal ultrasound (TRUS)-guided biopsy. For men presenting with MRI-visible disease, an MRI-US fusion-guided biopsy (via a transrectal or transperineal approach) was conducted; this involved obtaining two to four targeted cores per lesion, alongside 12 to 24 background systematic cores distributed across 12 anatomic sectors [1]. In scenarios involving diagnostic discordance between the initial histological findings and subsequent mpMRI, patients underwent a confirmatory targeted transperineal biopsy within a 3-month window. Patients whose disease was upgraded by this confirmatory sampling, thereby rendering them ineligible for AS under local protocols, were excluded from the analysis as they were deemed never to have formally commenced surveillance. While AS protocols inevitably vary between the two participating institutions, the standard follow-up schedule consisted of quarterly PSA testing combined with annual clinical reviews and MRI scans. Repeat histological sampling was either protocol-mandated at the 36-month mark or clinically triggered by three successive PSA elevations, marked radiological progression in lesion volume, or suspected stage progression on MRI [2].

#### Outcomes and exposures

The primary outcome was AS failure: a composite of oncological outcomes that indicate AS did not deliver its intended benefit of deferring treatment while maintaining expectation of cure with initial definitive therapy. These were: death from PCa, radiological progression to metastatic disease, histological progression to GG≥4 disease, or biochemical recurrence following curative-intent treatment (PSA ≥0.2 ng/mL after surgery; Phoenix definition after radiotherapy [3]).

To accurately characterise the safety profile of the AS protocol and mitigate confounding from downstream therapeutic outcomes, composite AS failures were further stratified according to the proximal index event that triggered AS exit. This hierarchical classification distinguished true surveillance failure from therapeutic failure and was defined as follows: i) *de novo* metastasis (radiological evidence of metastatic spread occurring as the initial failure event while the patient was still on AS), ii) progression to GG≥4 triggering AS exit (any subsequent BCR or metastasis occurring after definitive treatment was attributed to this initial index event), and iii) post-treatment BCR (isolated recurrence following curative-intent treatment for low-or intermediate-risk disease, occurring in the absence of prior GG≥4 progression or *de novo* metastasis and suggesting that clinically significant progression occurred during the surveillance period, ultimately compromising the window of curability).

All biopsies used in this study were retrospectively reviewed by expert genitourinary pathologists at two institutions to ensure definitive assessment of cribriform status, which may not have been reported consistently over the duration of this study. Pathologists at each participating site were blinded to patient outcome and used the International Society of Urological Pathology (ISUP) consensus definition [4] to classify patients as “cribriform-positive” if they had invasive cribriform carcinoma (ICC) and/or intraductal carcinoma (IDC). Since high-molecular-weight-keratin immunohistochemistry was not routinely available, IDC was diagnosed based on histopathological examination, defined as prostatic ducts filled up and distended by cribriform or solid carcinoma [5,6]. An example of a cribriform-positive tumour is provided in **Supplementary Figure S5**.

## Statistical analysis

Time-to-event analyses used the competing risk framework, in which discontinuation of AS in the absence of an AS failure event constitutes a clinically meaningful competing risk. For each patient, the outcome status variable was coded at three levels: (0) event-free and censored, defined as absence of AS failure criteria with no active treatment received at the last follow-up date; (1) AS failure event, as defined above; and (2) competing event, defined as discontinuation of AS with receipt of active treatment in the absence of AS failure criteria. Patients who progressed to GG≥4 disease without subsequently developing BCR or distant metastasis were censored at the date of first GG ≥4 or GG 5 detection, to preclude misattribution of outcome status in this subgroup.

Two multivariable Fine-Gray subdistribution hazard models were fitted to assess whether follow-up biopsy status and follow-up MRI status each predict AS failure independently of baseline risk. Both models used the same competing-risk structure as the primary cumulative incidence analyses and were adjusted for the same pre-specified baseline covariates (age, PSA density, grade group, and PI-RADS category). Critically, the two models were fitted on different analytic populations, because each exposure is defined only in the patients in whom it can be measured. The final MRI status model included all 619 men, as every patient underwent at least one follow-up MRI and therefore has a defined final MRI status. The final biopsy status model was necessarily restricted to the subset of men who underwent at least one repeat biopsy because cribriform status cannot be assigned without tissue sampling; men who never underwent repeat biopsy have an undefined (not negative) cribriform status and were therefore not classifiable on the exposure. Including them as a “cribriform-negative” reference would have misclassified unsampled patients as true negatives and biased the subdistribution hazard ratio upward.

These two models should therefore not be interpreted as a head-to-head comparison of the predictive performance of MRI versus biopsy. They differ in both their analytic population and the fact that one exposure (MRI) is universally measured while the other (biopsy) is conditional on having been sampled. Each model answers only whether its own exposure is an independent predictor of AS failure within the population in which that exposure is defined. The descriptive, whole-cohort comparison of radiological and histological predictors is provided by the cumulative incidence functions and the index-event distributions in, which include all enrolled patients and are not subject to this conditioning.

To mitigate immortal time bias, the time of origin for survival analyses was set to the date of the last available biopsy or MRI scan, not the date of initial AS enrolment. Specifically, for the final biopsy analysis, the time origin (T0) was the date of the last available follow-up biopsy. For patients classified as having stable disease without a recorded final biopsy date, T0 was set to the date of the first diagnostic (baseline) biopsy confirming AS eligibility. For the final MRI analysis, T0 was the date of the last available MRI scan. In both analyses, follow-up was accrued to the date of AS failure, date of last AS contact, or date of last documented follow-up, whichever occurred first. Rare cases in which T0 coincided with the end date were assigned a follow-up duration of 0.01 months to facilitate inclusion in cumulative incidence estimation.

The following percentages correspond to Sankey plots shown in **Fig. 2A-D**.

**Figure 2A**:

*MRI-invisible at baseline (n = 186):*

81% of lesions remained MRI-invisible at final MRI;

19% of patients developed MRI-visible lesions.

*MRI-visible at baseline (n = 433):*

85% of lesions remained MRI-visible at final MRI;

15% of patients no longer had MRI-visible lesions.

**Figure 2B**:

*MRI-invisible at baseline (n = 186):*

81% of lesions remained MRI-invisible;

17% of patients developed organ-confined MRI-visible lesions;

2% of patients developed locally advanced MRI-visible lesions.

*MRI-visible at baseline (n = 433):*

61% of lesions were radiologically stable or resolved;

32% of lesions increased in size or conspicuity;

6% of lesions showed definitive ≥rT3a progression;

1% of lesions progressed to metastatic disease.

Figure 2D:

*MRI-invisible at baseline (n = 186):*

36% of patients did not undergo repeat biopsy;

39% had GG1 disease on repeat biopsy;

12% progressed to NC GG2 disease;

2% progressed to NC GG3 disease;

1% progressed to NC ≥GG4 disease;

10% had cribriform disease detected on follow-up biopsy.

*MRI-visible at baseline (n = 433):*

50% of patients did not undergo repeat biopsy;

32% did not progress on repeat biopsy;

2% progressed to NC GG3 disease;

5% progressed to NC ≥GG4 disease;

9% had cribriform disease detected on follow-up biopsy.

Figure 2C:

*GG1 at baseline (n = 467):*

40% of patients did not undergo repeat biopsy;

46% did not progress on repeat biopsy;

7% progressed to NC GG2 disease;

1% progressed to NC GG3 disease;

1% progressed to NC ≥GG4 disease;

5% had cribriform disease detected on follow-up biopsy.

*GG2 at baseline (n = 152):*

37% of patients did not undergo repeat biopsy;

32% did not progress on repeat biopsy;

10% progressed to NC GG3 disease;

2% progressed to NC ≥GG4 disease;

2% of NC GG2 patients had cribriform detected on follow-up biopsy;

17% of GG2 patients remained or upgraded to cribriform ≥GG2.

---

**Supplementary Figure S1.**
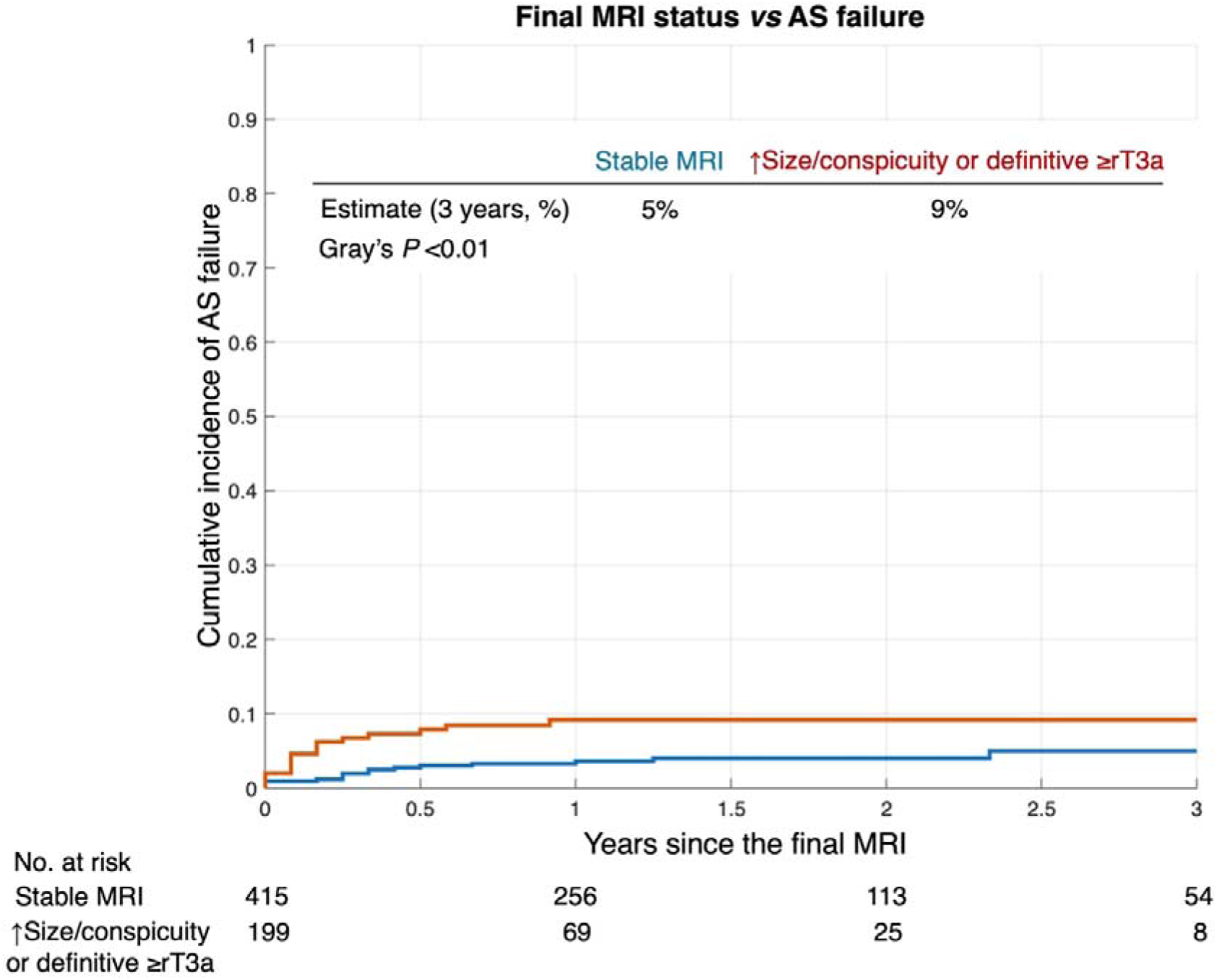
Supplementary competing-risk cumulative incidence analysis of AS failure stratified by final MRI status using a simplified two-group comparison. Patients with any radiological progression, defined as either a significant increase in lesion size or conspicuity or definitive progression to ≥rT3a disease, were combined into a single group and compared against those with radiologically stable disease. Patients with radiological N1/M1 disease were excluded from this comparison as they experienced AS failure by definition. Three-year cumulative incidence estimates were 5% for stable MRI and 9% for any MRI progression (Gray’s *P* <0.01). Numbers at risk represent patients remaining event-free and without a competing risk at each time point. AS, active surveillance; MRI, magnetic resonance imaging.

**Supplementary Figure S2.**
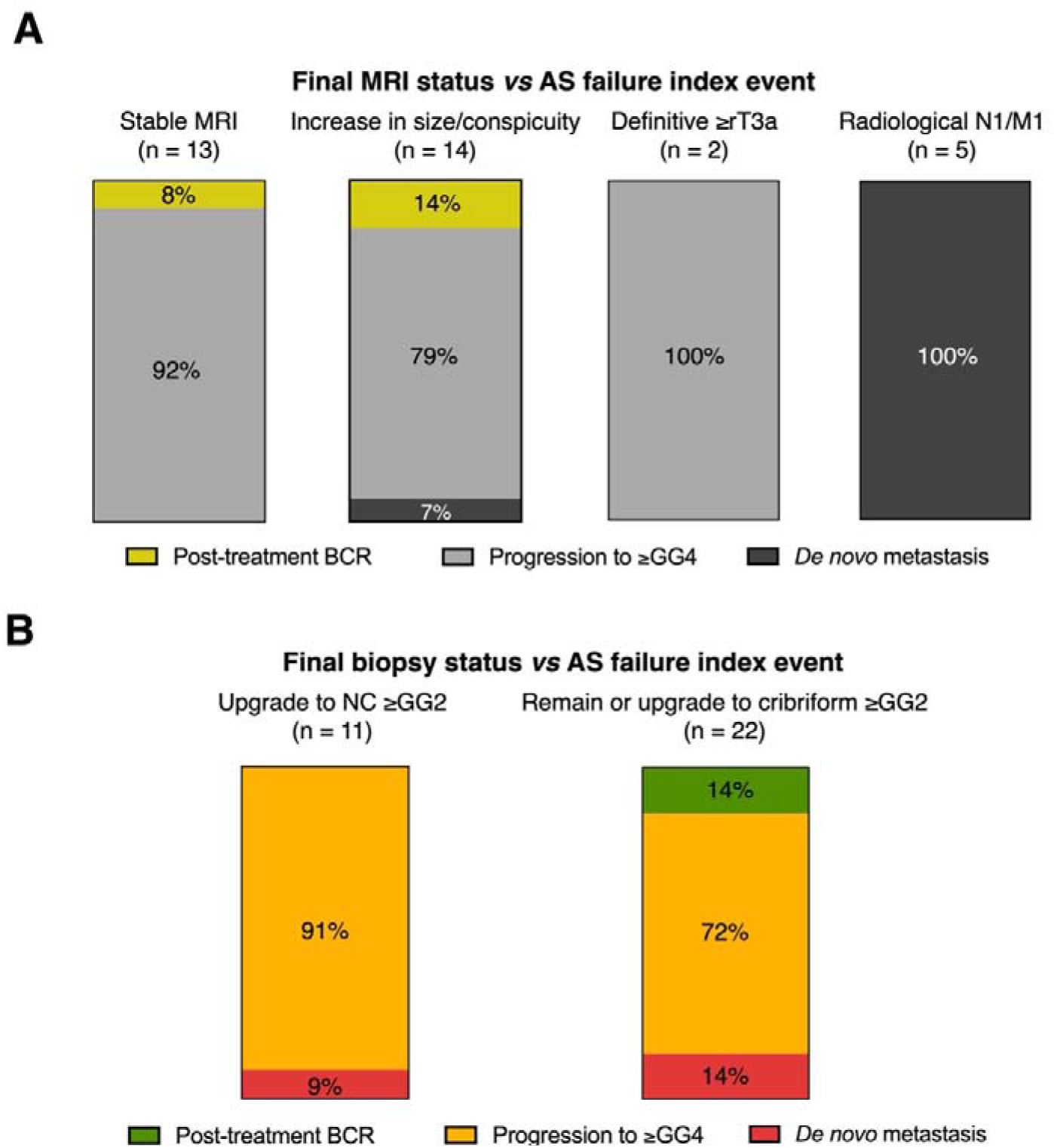
Distribution of AS failure index events stratified by final MRI and biopsy status category among patients who experienced AS failure. **(A)** Proportional distribution of AS failure index events across final MRI status categories. Among patients with a stable final MRI (n = 13), 92% had histological progression to GG≥4 as the index event and 8% had post-treatment BCR; none developed de novo metastasis. Among those with an increase in lesion size or conspicuity (n = 14), index events were histological progression to GG≥4 (79%), post-treatment BCR (14%), and de novo metastasis (7%). All patients with definitive ≥rT3a progression (n = 2) and radiological N1/M1 disease (n = 5) experienced GG ≥4 progression and de novo metastasis as their index events, respectively. **(B)** Proportional distribution of AS failure index events across final biopsy status categories, restricted to patients with non-cribriform GG≥2 upgrade (n = 11) and those with cribriform ≥GG2 morphology (n = 22) as their most adverse biopsy finding. Among NC ≥GG2 upgrade patients, 91% progressed through the GG ≥4 pathway and 9% had post-treatment BCR; none developed de novo metastasis. Among cribriform ≥GG2 patients, index events were histological progression to GG ≥4 (72%), post-treatment BCR (14%), and de novo metastasis (14%), reflecting a qualitatively more aggressive failure pattern consistent with the known biological aggressiveness of cribriform morphology. AS, active surveillance; BCR, biochemical recurrence; GG, grade group; MRI, magnetic resonance imaging; NC, non-cribriform.

**Supplementary Figure S3.**
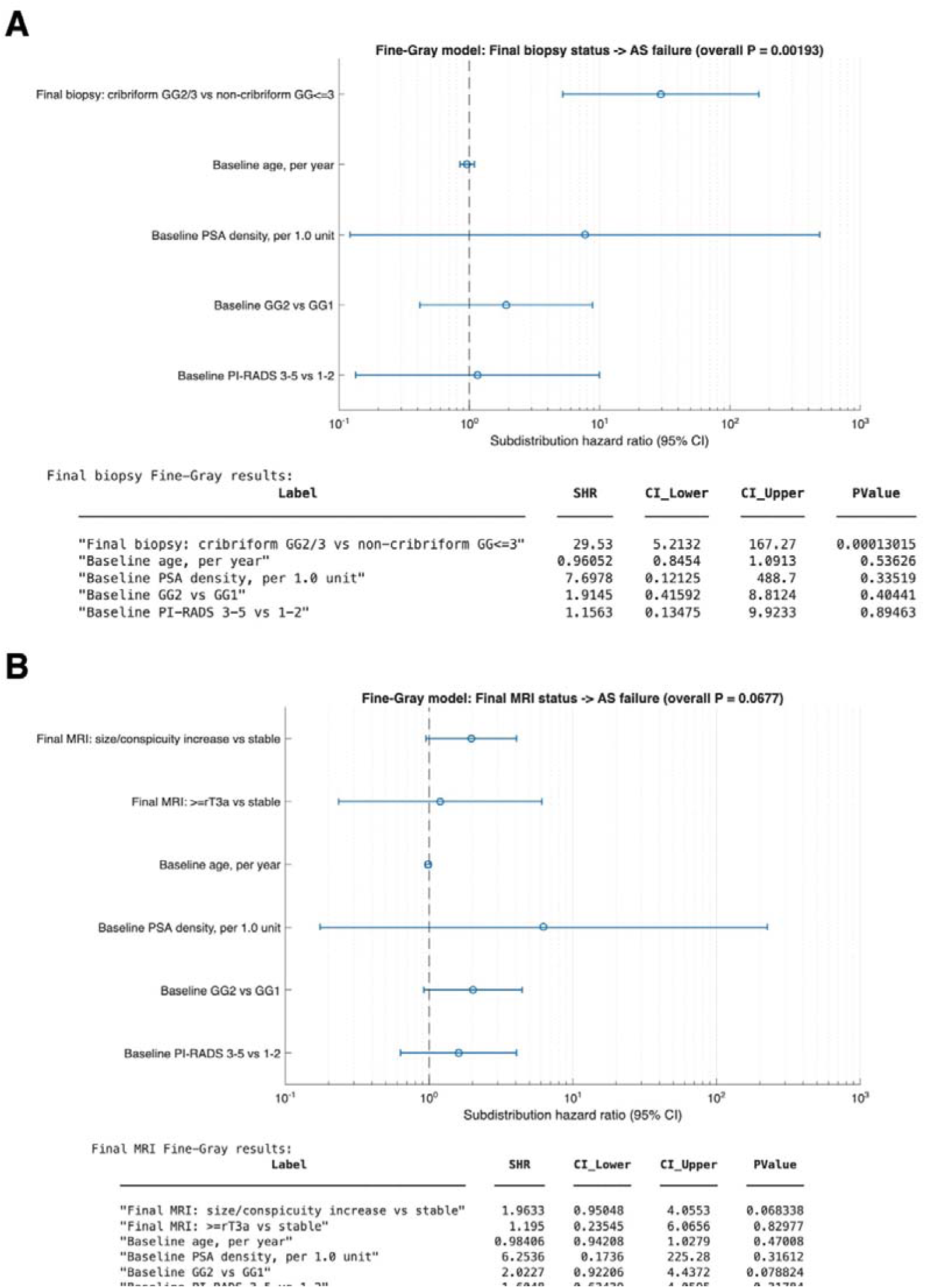
Multivariable Fine-Gray subdistribution hazard models for AS failure. **(A)** Final biopsy status model. **(B)** Final MRI status model. AS, active surveillance; CI, confidence interval; GG, grade group; MRI, magnetic resonance imaging; NC, non-cribriform; PI-RADS, Prostate Imaging-Reporting and Data System; PSA, prostate-specific antigen; SHR, subdistribution hazard ratio.

**Supplementary Figure S4.**
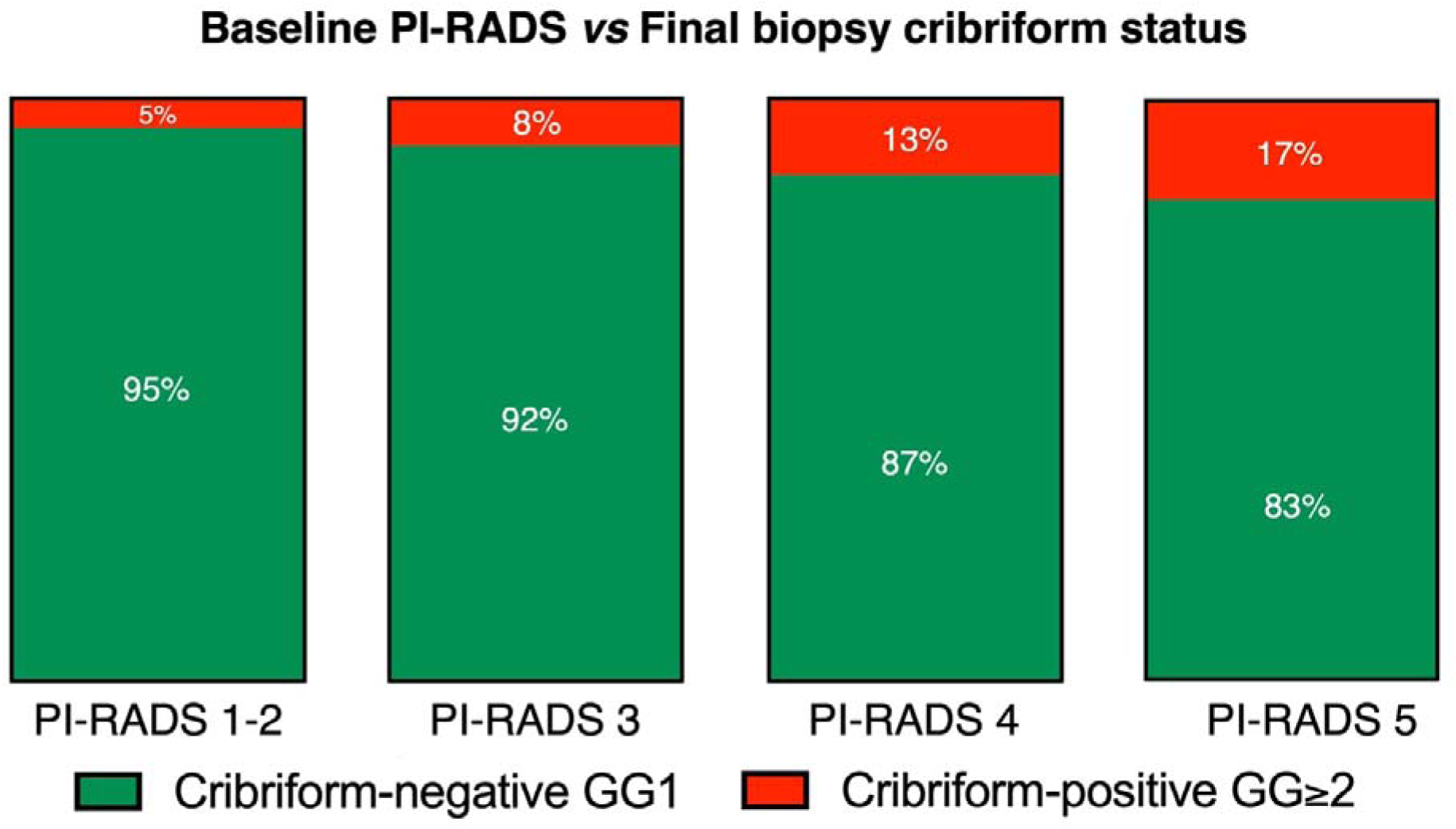
Proportional distribution of final biopsy cribriform status across baseline PI-RADS categories among 595 patients without cribriform. ≥**GG2 disease at baseline biopsy.** Each column represents 100% of patients within that PI-RADS category, subdivided by whether cribriform ≥GG2 disease was identified at any point during AS follow-up (red) or not (green). AS, active surveillance; GG, grade group; PI-RADS, Prostate Imaging-Reporting and Data System.

**Supplementary Figure S5.**
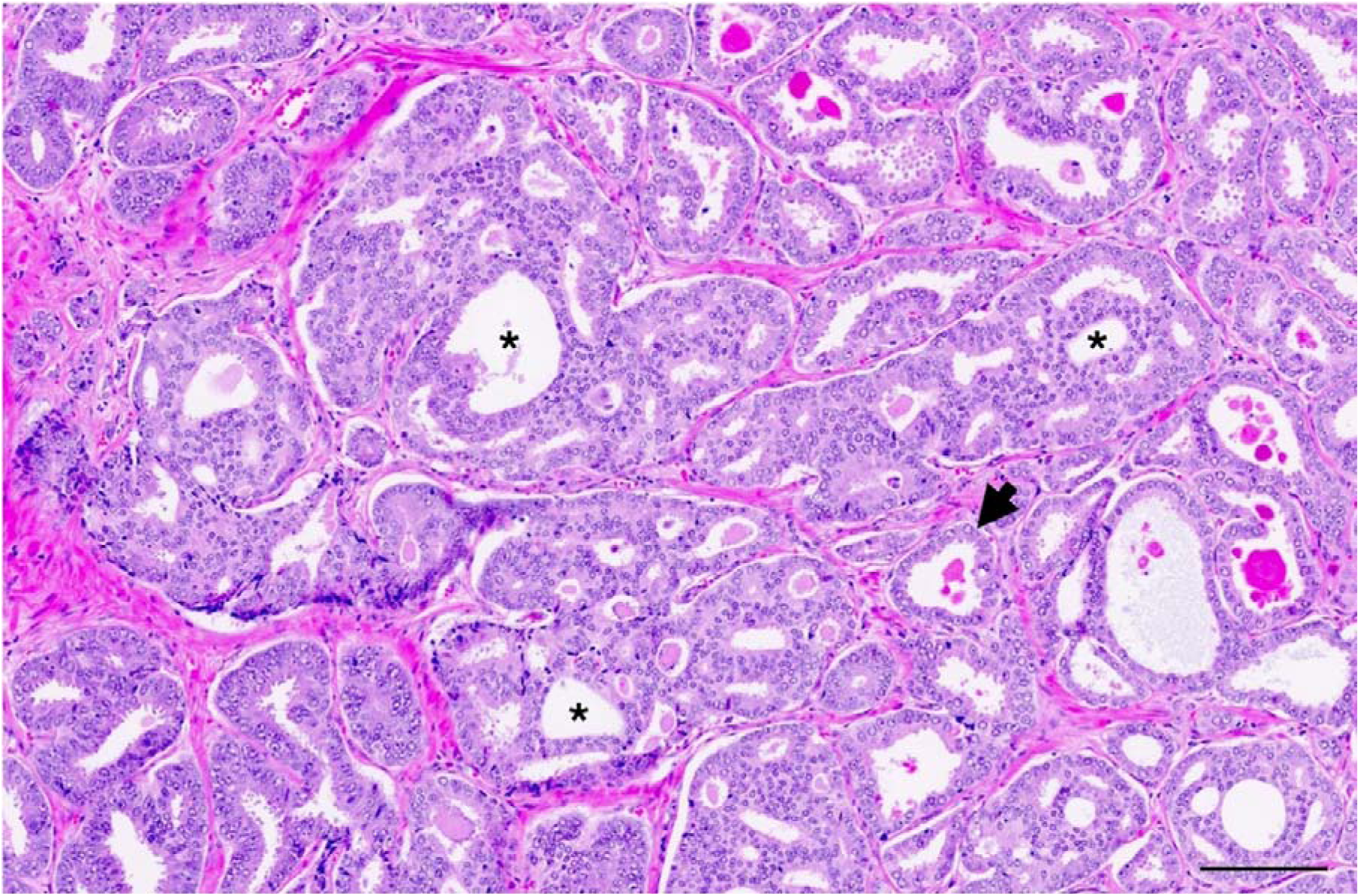
Representative H&E section of a biopsy sample from a patient who harboured cribriform-positive GG2 disease. Black asterisks denote individual glandular lumina within cribriform Gleason pattern 4 structures, defined as confluent sheets of contiguous malignant epithelial cells with multiple glandular lumina with no intervening stroma or mucin separating individual or fused glandular structures. A white arrow denotes an adjacent Gleason pattern 3 gland. Scale bar denotes 200 μm.

